# SARS-CoV-2 variants Omicron BA.4/5 and XBB.1.5 significantly escape T cell recognition in solid organ transplant recipients vaccinated against the ancestral strain

**DOI:** 10.1101/2023.08.14.23293991

**Authors:** Torin Halvorson, Sabine Ivison, Qing Huang, Gale Ladua, Demitra M. Yotis, Dhiraj Mannar, Sriram Subramaniam, Victor H. Ferreira, Deepali Kumar, Sara Belga, Megan K. Levings, the PREVenT study group

## Abstract

**Background:** Immune-suppressed solid organ transplant recipients (SOTRs) display impaired humoral responses to COVID-19 vaccination, but T cell responses are incompletely understood. The highly infectious SARS-CoV-2 variants Omicron BA.4/5 and XBB.1.5 escape neutralization by antibodies induced by vaccination or infection with earlier strains, but T cell recognition of these lineages in SOTRs is unclear.

**Methods:** We characterized Spike-specific T cell responses to ancestral SARS-CoV-2, Omicron BA.4/5 and XBB.1.5 peptides in a prospective study of kidney, lung and liver transplant recipients (n = 42) throughout a three- or four-dose ancestral Spike mRNA vaccination schedule. Using an optimized activation-induced marker assay, we quantified circulating Spike-specific CD4+ and CD8+ T cells based on antigen-stimulated expression of CD134, CD69, CD25, CD137 and/or CD107a.

**Results:** Vaccination strongly induced SARS-CoV-2-specific T cells, including BA.4/5- and XBB.1.5-reactive T cells, which remained detectable over time and further increased following a fourth dose. However, responses to Omicron BA.4/5 and XBB.1.5 were significantly lower in magnitude compared to ancestral strain responses. Antigen-specific CD4+ T cell frequencies correlated with anti-receptor-binding domain (RBD) antibody titres, with post-second dose T cell responses predicting subsequent antibody responses. Patients receiving prednisone, lung transplant recipients and older adults displayed weaker responses.

**Conclusions:** Ancestral strain vaccination stimulates BA.4/5 and XBB.1.5-cross-reactive T cells in SOTRs, but responses to these variants are diminished. Antigen-specific T cells can predict future antibody responses and identify vaccine responses in seronegative individuals. Our data support monitoring both humoral and cellular immunity in SOTRs to track effectiveness of COVID-19 vaccines against emerging variants.

## Introduction

COVID-19 vaccines effectively reduce infections, hospitalizations and mortality from SARS-CoV-2^1–3^. However, immune-suppressed populations such as solid-organ transplant recipients (SOTRs), who require lifelong immune-suppressive therapy to prevent allograft rejection, remain at elevated risk from COVID-19 despite vaccination^4^. SOTRs display impaired humoral immunity to COVID-19 vaccination, with decreased anti-Spike binding and neutralizing antibody (nAb) responses compared to the general population^5–8^. However, a third or fourth ‘booster’ dose significantly increases seropositivity and nAbs in SOTRs^6,9,10^.

As SARS-CoV-2 continues to evolve, emerging variants, including Omicron lineages and their derivatives, continue to challenge global COVID-19 immunity^11–15^. The Omicron BA.4 and BA.5 (henceforth BA.4/5) subvariants display over 40 Spike protein mutations and enhanced affinity for the ACE2 receptor^13,16^. More recently, recombinant BA.2-derived XBB lineages have emerged, with XBB.1.5 the latest to dominate globally^14,15,17,18^. These subvariants are highly infectious and evade neutralization by vaccination- or infection-induced nAbs targeting the ancestral strain^16^. Neutralization of XBB lineages is reduced more than 100-fold relative to ancestral SARS-CoV-2, even after vaccination with BA.4/5 bivalent vaccines^17–20^.

Cellular immunity to SARS-CoV-2 in SOTRs is less well understood. SARS-CoV-2-specific T cell responses are widely recognized to be critical in mitigating severe COVID-19 disease^21–23^, with CD4+ and CD8+ responses correlating negatively with disease severity^24^ and breakthrough infection risk post-vaccination^25^. Furthermore, memory T cells cross-recognize emerging variants, including Omicron and the BA sublineages^18,25–27^. However, SOTRs display poor cellular responses to vaccination, with lower Spike-specific CD4+ and CD8+ T cell frequencies compared to healthy controls two weeks and six months after two-dose vaccination^25,28^. Vaccination also induces comparatively weaker Spike-specific IFN-γ+ and/or IL-2+ T cell responses than natural infection in SOTRs^29,30^.

While it is well-established that BA.4/5 and XBB.1.5 escape antibody neutralization, whether vaccination against the ancestral strain induces T cell responses cross-recognizing these variants is unclear in SOTRs. A recent study assessed T cell responses to a fourth mRNA vaccine dose by intracellular cytokine staining (ICS) in a small cohort of SOTRs, finding that a fourth dose significantly increased frequencies of IFN-γ+/IL-2+ BA.4/5-specific CD4+, but not CD8+, T cells^9^. Only one study has directly compared ancestral SARS-CoV-2- and BA.4/5-specific T cell responses in SOTRs, showing no differences in magnitude by ICS^31^. However, ICS does not quantify the full diversity of antigen-specific T cells^32,33^. Alternatively, activation-induced marker (AIM) assays leverage T cell surface protein upregulation in response to antigen-specific stimulation, broadly quantifying antigen-specific T cells independently of proliferation or cytokine production^32–35^. Indeed, AIM assays are increasingly used to assess T cell responses to SARS-CoV-2 infection and vaccination^24,27,28,36^.

Here, we used the AIM method to characterize vaccine-induced SARS-CoV-2-specific T cells in a prospective cohort of 42 liver, kidney and lung transplant recipients. Of specific interest was whether vaccination against the SARS-CoV-2 ancestral strain would stimulate T cells with cross-reactivity to Omicron BA.4/5 and/or XBB.1.5.

## Methods (see also Extended Methods)

### Study Design and Enrollment

The PREVenT-COVID study is a Canadian prospective study of vaccine immunogenicity in SOTRs across seven tertiary care transplant centres^9,37^. This study of cell-mediated immunity was conducted at the University of British Columbia (UBC) with approval from the UBC Research Ethics Board (H21-01269). Adult SOTRs provided informed consent and were enrolled at first dose of an approved monovalent COVID-19 mRNA vaccine, BNT162b2 (Pfizer-BioNTech,) or mRNA-1273 (Moderna), beginning January 2021. Whole blood was drawn from participants within 2 weeks prior to the second dose, 3-6 weeks post-second dose, 6 months post-first dose, 3-6 weeks post-third dose and one year post-first dose. One-year samples served as post-fourth dose (3-6 weeks) samples in a subset of patients. PBMCs were collected from blood samples and cryopreserved at -80°C. Participants were to inform the study team if they developed COVID-19 for confirmation via PCR or rapid antigen test.

### Activation-Induced Marker Assays

SARS-CoV-2 ancestral strain (PM-WCPV-S-1, JPT) and Omicron BA.4/5 (PM-SARS2-SMUT10-1, JPT) overlapping peptide pools corresponding to the complete Spike proteins were aliquoted in 30% DMSO and stored at -80°C. Cryopreserved PBMCs were thawed into 37°C Immunocult XF (10981, STEMCELL) with 1% penicillin/streptomycin and 50 U/mL benzonase (70664-3, Novagen). PBMCs were rested overnight at 37°C in Immunocult XF with 1% penicillin/streptomycin and stimulated for 20 h at 4-5×10^5^ cells per condition in 200 uL with 1 µg/mL SARS-CoV-2 ancestral strain or BA.4/5 Spike peptides, equimolar dimethyl sulfoxide (DMSO, 0.12%), Fluzone® Quadrivalent influenza vaccine (2.5%) (Sanofi-Pasteur) or Cytostim (0.05%) (130-092-172, Miltenyi). CD4 and CD107a staining antibodies were added during stimulation, with surface staining for CD3, CD4, CD8, CD25, CD69, CD134, and CD137 after stimulation. Data were acquired on an LSR Fortessa (BD) flow cytometer on high-throughput sampler (HTS) mode. Antigen-specific CD4+ T cells were defined as CD134+/CD25+, CD134+/CD69+ or CD137+/CD69+ while antigen-specific CD8+ T cells were defined as CD107a+/CD69+, CD107a+/CD137+ or CD137+/CD69+, as previously described^24,38–46^.

Whole protein stimulations were performed with protein isolates (**Extended Methods**) from ancestral SARS-CoV-2 or XBB1.5 at 0.5 µg/mL, equivolumetric PBS, 1 µg/mL ancestral strain peptides or 0.05% Cytostim for 44 h.

### Data Analysis

Data were analyzed using FlowJo v10.8.1 and GraphPad Prism v10.0.0. Antigen-specific T cells were quantified as the percentage of CD4+ or CD8+ T cells expressing each AIM marker, after subtracting the frequency of AIM+ cells in the equivalent unstimulated condition. Net AIM frequencies less than 0.005%, considered the limit of detection for the assay, were set to equal 0.005% to avoid negative or zero values. Samples with fewer than 10 000 total CD4+ or CD8+ T cells were excluded. AIM responses were compared across timepoints using a mixed-effects analysis (REML) on log2-transformed data, assuming sphericity, with post-hoc Dunnett’s multiple comparisons test. Patients who received a fourth dose or contracted COVID-19 were initially excluded to assess the duration of vaccine-induced T cell immunity.

Responses at one year were compared within (paired Wilcoxon tests) and between (Kruskal-Wallis test with Dunn’s multiple comparisons) control, post-fourth dose (within 3-6 weeks) and hybrid (fourth dose and contracted COVID-19) groups. Post-third dose BA.4/5 and XBB.1.5 responses were compared to the ancestral strain by paired Wilcoxon tests. Post-third dose AIM responses between groups defined by clinical parameters were compared using Mann-Whitney and Kruskal-Wallis tests as appropriate. Semilogarithmic regression analysis modelled the continuous effects of age and time since transplantation on post-third dose AIM responses.

Spearman’s correlations assessed relationships between post-third dose T cell AIM responses and published serum anti-RBD titres from the same individuals^9^. In patients with undetectable anti-RBD antibodies post-second dose, log-log regression modelled the relationship between post-second dose CD4+ AIM responses and post-third dose anti-RBD antibodies. Post-third dose BA.4/5-specific AIM responses were compared between individuals with and without detectable BA.4/5-neutralizing antibodies^9^ by paired t-tests on log2-transformed AIM+ frequencies.

## Results

### Study Population

The study cohort consisted of 42 SOTRs with a median age of 58 (IQR 47-56), of which 22 (52%) were female and 20 (48%) were male (**Table 1, Figure S1**). Subjects were recipients of kidney (n = 16), liver (n = 16) or lung (n = 10) transplants. Median time since transplantation was 6.8 years (IQR 3.1-12.9). All subjects received at least two doses of monovalent Pfizer-BioNTech BNT162b2 or Moderna mRNA-1273 COVID-19 vaccines. As one-year samples were collected soon (3-6 weeks) after patients received a fourth dose, subjects were grouped as neither having contracted COVID-19 nor received a fourth dose (controls, n = 6), post-fourth dose (n = 15), post-fourth dose and contracted COVID-19 (‘hybrid’, n = 7) or contracted COVID-19 only (n = 4). At enrollment, all patients were receiving at least one immune-suppressive drug, with tacrolimus (35/42, 83%) mycophenolate mofetil or mycophenolate sodium (27/42, 64%) and prednisone (20/42, 48%) being the most frequent.

**Table 1.** Study Cohort Characteristics.

| Characteristic | n (%) or median [IQR]<sup>a</sup> |  |  | Total |
| --- | --- | --- | --- | --- |
|  | Kidney | Liver | Lung |  |
| *Sample size* | 16 | 16 | 10 | 42 |
| Pre-2nd dose | 5 (31%) | 12 (75%) | 4 (40%) | 21 (50%) |
| Post-2nd dose | 16 (100%) | 16 (100%) | 8 (80%) | 40 (95%) |
| Six months post-1st dose | 3 (19%) | 4 (25%) | 4 (40%) | 11 (26%) |
| Post-3rd dose | 14 (88%) | 11 (69%) | 8 (80%) | 33 (79%) |
| One year post-1st dose | 14 (88%) | 9 (56%) | 9 (90%) | 32 (76%) |
| Neither 4th dose nor COVID-19 | 1 (6%) | 4 (25%) | 1 (10%) | 6 (14%) |
| Post-4th dose (no COVID-19) | 7 (44%) | 4 (25%) | 4 (40%) | 15 (36%) |
| Hybrid (4th dose + COVID-19) | 6 (38%) | 1 (6%) | 0 (0%) | 7 (17%) |
| COVID-19 (no 4th dose) | 0 (0%) | 0 (0%) | 4 (40%) | 4 (10%) |
| *Sample collection timing* |  |  |  |  |
| 1st dose to pre-2nd dose sample (days) | 73<br>[40, 84] | 71<br>[47, 76] | 70<br>[69, 75] | 70<br>[53, 79] |
| 2nd dose to post-2nd dose sample (days) | 29<br>[26, 33] | 29<br>[28, 33] | 34<br>[29, 35] | 29<br>[26, 34] |
| 2nd dose to 6 month sample (days) | 105 | 146<br>[128, 152] | 105<br>[102, 108] | 109<br>[100, 134] |
| 3rd dose to post-3rd dose sample (days) | 33<br>[28, 40] | 31<br>[29, 36] | 32<br>[28, 36] | 32<br>[28, 39] |
| 3rd dose to 1 year sample (no 4th dose or COVID-19) (days) | 232 | 203<br>[194, 209] | 217 | 211<br>[200, 216] |
| 4th dose to 1 year sample (no COVID-19) (days) | 28<br>[27, 39] | 22<br>[20, 27] | 30<br>[25, 35] | 27<br>[22, 39] |
| 4th dose to 1 year sample (hybrid) (days) | 34<br>[26, 39] | 36 |  | 35<br>[28, 38] |
| COVID-19 to 1 year sample (hybrid) (days) | 135<br>[110, 151] | 118 |  | 126<br>[111, 148] |
| *Age* | 55<br>[46, 66] | 62<br>[50, 66] | 56<br>[49, 65] | 58<br>[47, 66] |
| *Time since transplant (y)* | 4.6<br>[2.8, 8.0] | 8.7<br>[4.5, 13.9] | 9.0<br>[4.3, 14.1] | 6.8<br>[3.2, 12.9] |
| *Sex* |  |  |  |  |
| Female | 8 (50%) | 9 (56%) | 5 (50%) | 22 (52%) |
| Male | 8 (50%) | 7 (44%) | 5 (50%) | 20 (48%) |
| *Vaccinations* |  |  |  |  |
| 1st dose | 16 (100%) | 16 (100%) | 10 (100%) | 42 (100%) |
| Moderna | 2 (13%) | 6 (38%) | 0 (0%) | 8 (19%) |
| Pfizer-BioNTech | 14 (88%) | 10 (63%) | 10 (100%) | 34 (81%) |
| 2nd dose | 16 (100%) | 16 (100%) | 10 (100%) | 42 (100%) |
| Moderna | 2 (13%) | 8 (50%) | 0 (0%) | 10 (24%) |
| Pfizer-BioNTech | 14 (88%) | 8 (50%) | 10 (100%) | 32 (76%) |
| 3rd dose | 15 (94%) | 13 (81%) | 10 (100%) | 38 (90%) |
| Moderna | 6 (38%) | 11 (69%) | 6 (60%) | 23 (55%) |
| Pfizer-BioNTech | 9 (56%) | 2 (13%) | 4 (40%) | 15 (36%) |
| 4th dose | 14 (88%) | 12 (75%) | 9 (90%) | 35 (83%) |
| Moderna | 11 (69%) | 11 (69%) | 7 (70%) | 29 (69%) |
| Pfizer-BioNTech | 3 (19%) | 1 (6%) | 2 (20%) | 6 (14%) |
| *Immunosuppression at enrollment* |  |  |  |  |
| Prednisone | 10 (63%) | 0 (0%) | 10 (100%) | 20 (48%) |
| Cyclosporine | 0 (0%) | 1 (6%) | 0 (0%) | 1 (2%) |
| Tacrolimus | 16 (69%) | 11 (69%) | 10 (100%) | 35 (83%) |
| Mycophenolate mofetil or mycophenolate sodium | 12 (44%) | 7 (44%) | 8 (80%) | 27 (64%) |
| Azathioprine | 3 (19%) | 1 (6%) | 1 (10%) | 5 (12%) |
| Sirolimus | 0 (0%) | 3 (19%) | 0 (0%) | 3 (7%) |

<sup>a</sup>Only medians are reported when groups have fewer than n = 4

### COVID-19 vaccination induces significant and durable BA.4/5-specific T cell responses in solid-organ transplant recipients

We assessed T cell AIM responses following stimulation with SARS-CoV-2 ancestral strain or BA.4/5 peptides throughout a 3-4 dose COVID-19 vaccination schedule (**Figure 1A**). Antigen-specific CD4+ T cells (CD134+/CD25+, CD134+/CD69+ or CD137+/CD69+) and CD8+ T cells (CD107a+/CD69+, CD107a+/CD137+ or CD137+/CD69+) were quantified (**Figure 1B, Figure S2**). Ancestral- and BA.4/5-specific CD4+ AIM responses increased significantly after a second dose (p <0.0001) but trended lower at six months (**Figure 1C**). A third dose induced significant responses that remained elevated above pre-second dose levels at one year (p <0.0001). Similar results were observed for CD8+ T cell responses, but with lower AIM+ frequencies and substantial heterogeneity between donors (**Figure 1C**). However, post-third dose responses to ancestral SARS-CoV-2 were significant for all CD8 AIM markers analyzed, and BA.4/5-specific CD137+/CD69+ responses were significant at post-second dose, post-third dose and one-year timepoints.

**Figure 1.**
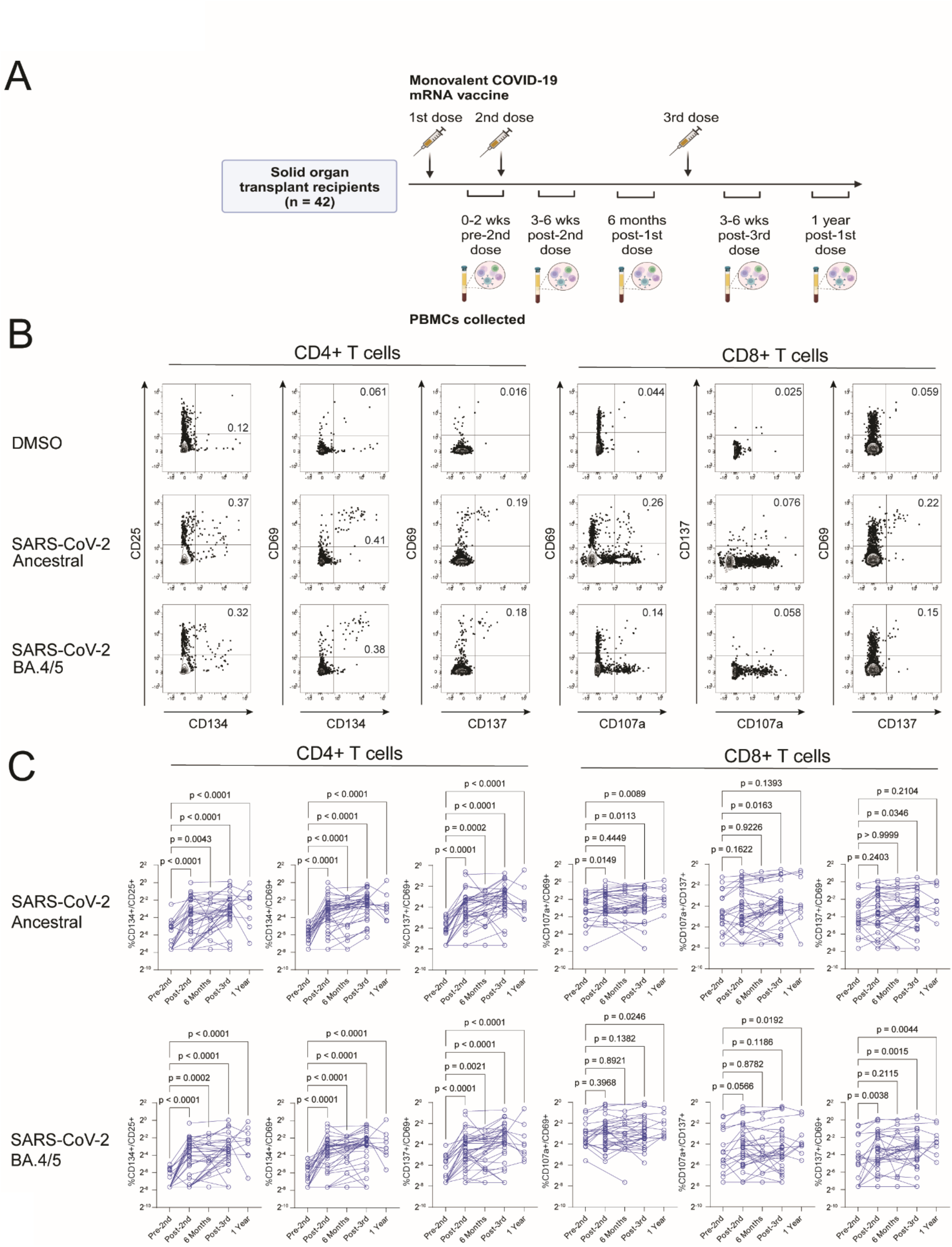
Second and third vaccine doses induce durable Omicron BA.4/5-specific T cell responses in solid-organ transplant recipients. A) PBMC samples from solid organ transplant recipients (SOTRs, n = 42) immunized with COVID-19 mRNA vaccines were collected at pre-second dose, post-second dose, six months post-first dose, post-third dose and one-year timepoints. B) Representative flow cytometry data showing T cell activation-induced marker (AIM) responses to the ancestral strain of SARS-CoV-2 in one donor after three doses of a COVID-19 mRNA vaccine. Samples were stimulated with 1 µg/mL SARS-CoV-2 ancestral strain or Omicron BA.4/5 Spike peptides for 20 h. CD4+ T cell activation-induced marker (AIM) responses were measured as frequencies of CD134+/CD25+, CD134+/CD69+ or CD137+/CD69+ events among CD4+ T cells. CD8+ AIM responses were measured as frequencies of CD107a+/CD69+, CD107a+/CD137+ or CD137+/CD69+ events among CD8+ T cells. All data represent net AIM+ frequencies after subtracting the equivalent AIM+ frequency in the DMSO-stimulated control. C) Time course of CD4+ and CD8+ T cell AIM responses to ancestral SARS-CoV-2 and BA.4/5 in SOTRs. Participants who had already received a third dose were excluded at the six-month timepoint, and participants who received a fourth dose or developed COVID-19 were excluded at the one-year timepoint to model the natural history of the vaccine-induced T cell response. P-values represent Dunnett’s multiple comparisons test following a mixed effects analysis on log2-transformed AIM+ frequencies.

As a control, we measured CD4+ and CD8+ responses to an inactivated influenza vaccine (Fluzone® Quadrivalent, Sanofi-Pasteur) at the same timepoints (**Figure S3A, B, C**). COVID-19 vaccination induced no changes in influenza-specific T cell responses, supporting the specificity of our assays. Furthermore, cryopreserved replicate PBMC aliquots from two healthy controls showed stable AIM responses over several months (**Figure S4**), demonstrating low technical variation of the assay. For simplicity, we focused further analyses on CD134+/CD69+ CD4+ T cell responses, and CD137+/CD69+ CD8+ T cell responses, as these AIMs are widely used^24,33,44,45,47–49^ and showed the strongest trends across vaccination timepoints.

### A fourth monovalent ancestral vaccine dose boosts BA.4/5-specific T cell responses

A subset of patients received a fourth dose 3-6 weeks prior to one-year sample collection, with some additionally contracting COVID-19 (hereafter ‘hybrid’) (**Figure 2A**), allowing us to ask if a 4^th^ dose further increased BA.4/5-specific T cell immunity. Indeed, fourth dose recipients and hybrid patients showed trending or significant increases above post-third dose levels in ancestral- and BA.4/5-specific CD4+ responses (**Figure 2B**), with these increases being significantly greater than observed in controls who received no fourth dose (**Figure 2C**). No significant differences were identified between the fourth dose and hybrid groups.

**Figure 2.**
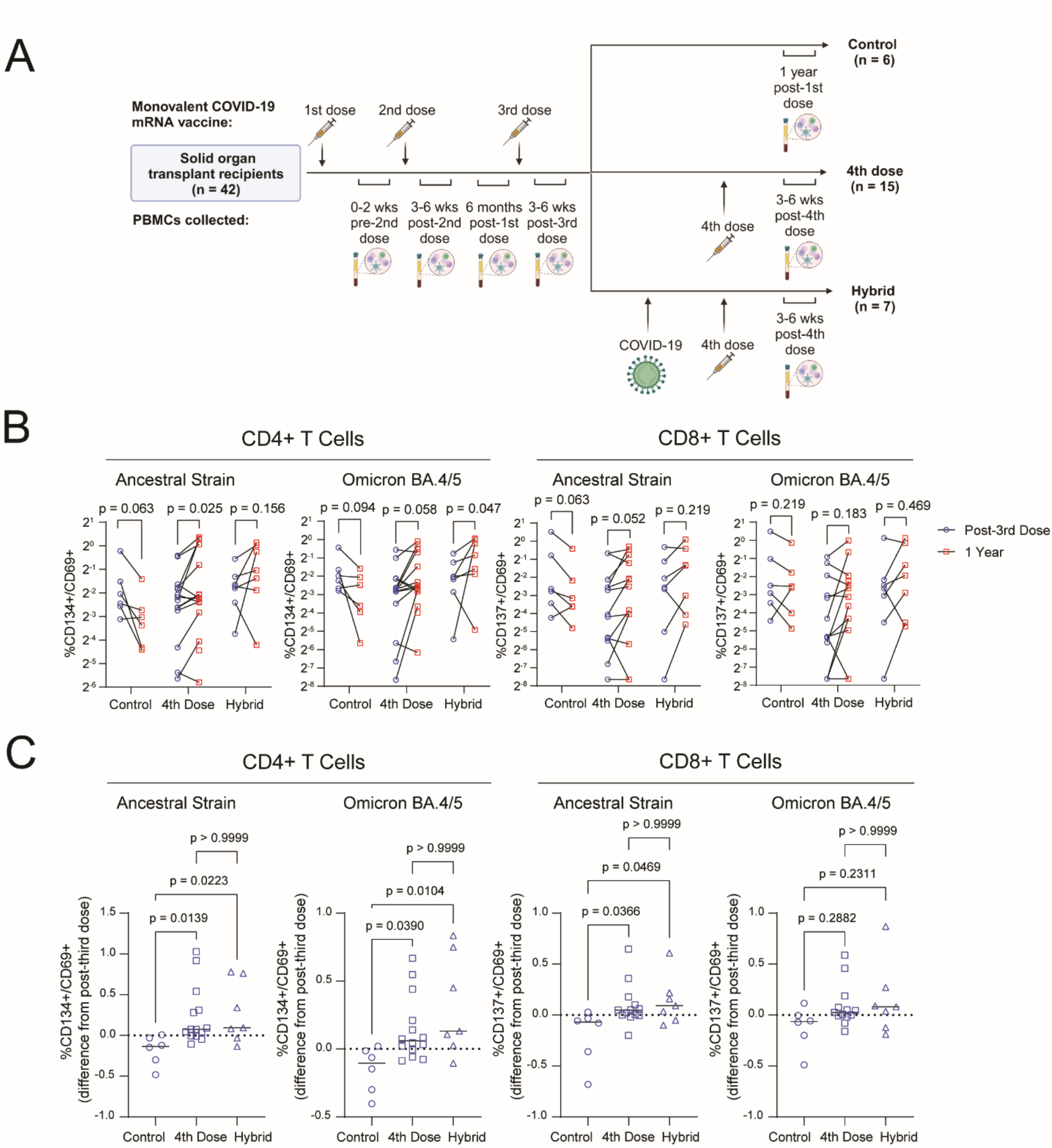
A fourth vaccine dose enhances SARS-CoV-2-specific T cell responses in solid organ transplant recipients. PBMCs from solid organ transplant recipients (SOTRs) were collected at post-third dose and one year post-first dose (6-8 months post-third dose) timepoints and stimulated with 1 µg/mL SARS-CoV-2 ancestral strain or Omicron BA.4/5 Spike peptides for 20 h. T cell activation-induced marker (AIM) responses were quantified as CD134+/CD69+ or CD137+/CD69+ frequencies among CD4+ and CD8+ T cells, respectively, after subtraction of the equivalent AIM+ frequency in the DMSO-stimulated control. **A)** At the one-year timepoint, SOTRs were grouped as having received neither a fourth dose nor a contracting COVID-19 (controls, n = 6), having received a fourth monovalent COVID-19 mRNA vaccine dose within 3-6 weeks of sample collection (fourth dose, n = 15), or a fourth dose and symptomatic COVID-19 (hybrid, n = 7). Patients who had only COVID-19 infection (n = 4) were excluded due to insufficient sample size. **B)** Evolution of CD4+ and CD8+ T cell AIM responses in from the post-third dose to one-year timepoint within fourth dose, hybrid and control groups of SOTRs. Paired Wilcoxon signed-rank test p-values are shown. **C)** Effect of a fourth dose or hybrid immunity on CD4+ and CD8+ T cell AIM responses in SOTRs. Changes from post-third dose to one-year timepoints are compared between fourth dose, hybrid and control groups. P-values represent Dunn’s multiple comparisons test following a Kruskal-Wallis test.

*T cell responses to Omicron BA.4/5 are lower in magnitude than responses to the ancestral strain*

Having established that second and third doses of the monovalent ancestral vaccine induce BA.4/5-responsive CD4+ and CD8+ T cells, we next compared the magnitude of post-third dose BA.4/5-specific AIM responses with ancestral strain responses. Among CD4+ T cells, responses to peptides from Omicron BA.4/5 were lower compared to ancestral SARS-CoV-2: median frequencies of CD134+/CD25+ (1.67-fold lower, Wilcoxon signed-rank test p <0.0001), CD134+/CD69+ (1.38-fold lower, p <0.0001), and CD137+/CD69+ (1.34-fold lower, p = 0.0022) were diminished (**Figure 3A**). Similar trends were observed among CD8+ T cells, particularly when comparing CD137+/CD69+ frequencies (1.45-fold lower, p = 0.0071) (**Figure 3B**).

**Figure 3.**
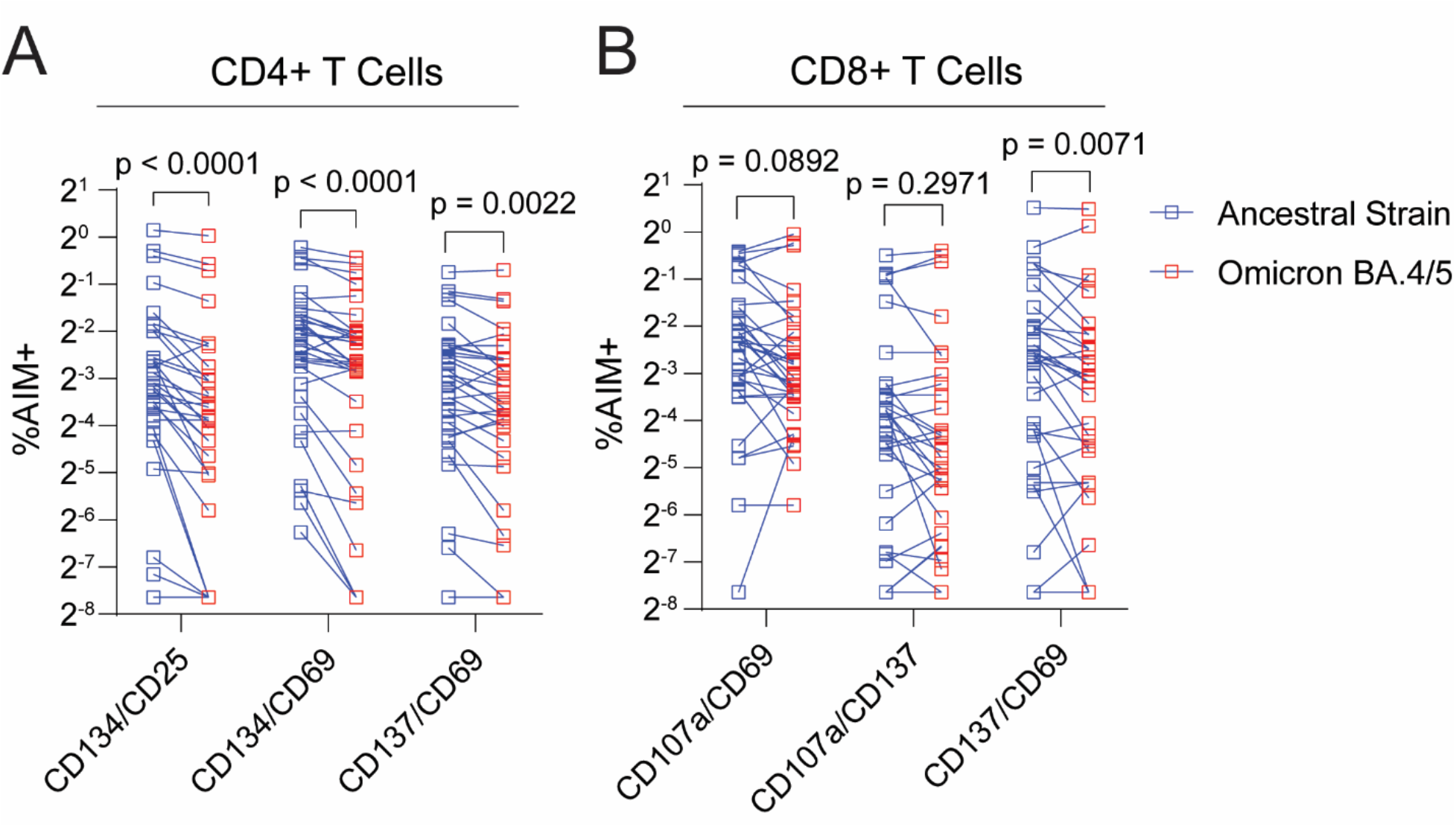
T cell activation-induced marker responses to Omicron BA.4/5 are weaker than responses to ancestral SARS-CoV-2 in solid organ transplant recipients. Paired comparisons of CD4+ (**A**) and CD8+ (**B**) T cell AIM responses to Omicron BA.4/5 and ancestral SARS-CoV-2, measured after three doses of a COVID-19 mRNA vaccine, in solid organ transplant recipients (n = 33). Following a 20-h stimulation of patient PBMCs with 1 µg/mL SARS-CoV-2 ancestral strain or Omicron BA.4/5 Spike peptides, CD4+ T cell activation-induced marker (AIM) responses were quantified as frequencies of CD134+/CD25+, CD134+/CD69+ or CD137+/CD69+ events among CD4+ T cells, while CD8+ AIM responses were measured as frequencies of CD107a+/CD69+, CD107a+/CD137+ or CD137+/CD69+ events among CD8+ T cells, after subtracting the equivalent AIM+ frequency in the DMSO-stimulated control. Wilcoxon signed-rank test p-values are shown.

### Patients receiving prednisone, lung transplant recipients and older individuals display weaker T cell responses

We next sought to identify patient characteristics associated with weaker BA.4/5-specific AIM responses, using post-third dose data for its large sample size and clinical relevance. Compared to patients not receiving prednisone, patients on prednisone at baseline showed significantly lower CD4+ (1.53-fold lower, Mann-Whitney U-test p = 0.030) and CD8+ (3.36-fold lower, p = 0.017) responses to BA.4/5, with similar trends for ancestral strain responses (**Figure 4A**). Since all lung and no liver recipients were receiving prednisone at baseline, we also compared responses by organ type (**Figure 4B**), showing trending or significantly lower BA.4/5-specific CD4+ AIM responses in lung transplant recipients compared to liver (4.97-fold lower, p = 0.0017) and kidney (3.23-fold lower, p = 0.0877), with similar results for ancestral responses (lung vs liver, 3.28-fold lower, p = 0.0029; lung vs kidney, 3.71-fold lower, p = 0.0161) (**Figure 4B**). CD8+ AIM responses followed similar trends but were non-significant.

**Figure 4.**
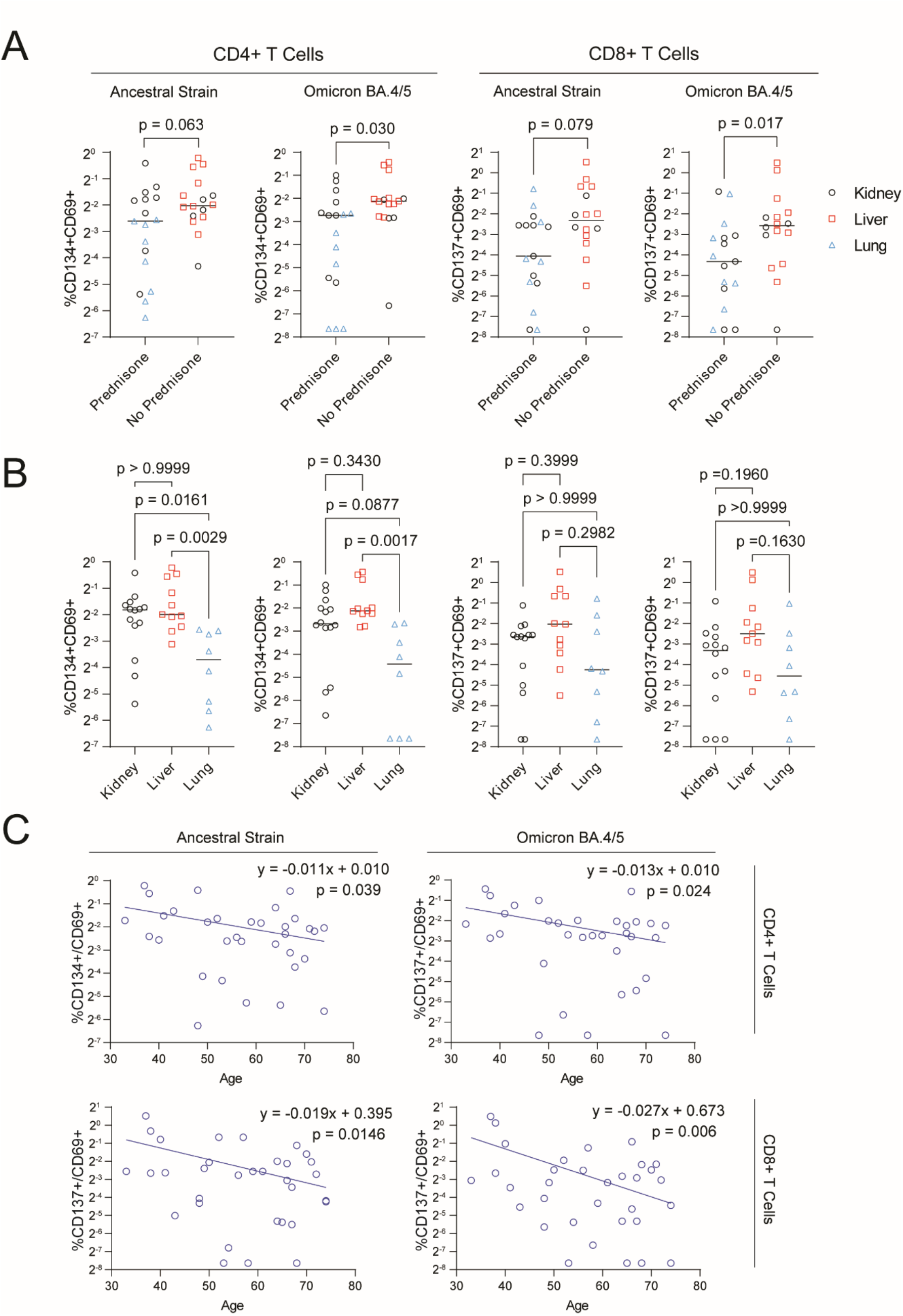
Prednisone, lung transplantation and older age are associated with weaker T cell responses to COVID-19 vaccination. Post-third dose CD4+ and CD8+ T cell activation-induced marker (AIM) responses to SARS-CoV-2 ancestral and Omicron BA.4/5 variants are shown in solid-organ transplant recipients. Patient PBMCs were stimulated for 20 h with 1 µg/mL SARS-CoV-2 ancestral strain or Omicron BA.4/5 Spike peptides, and T cell activation-induced marker responses were quantified by flow cytometry as frequencies of CD134+/CD69+ events among CD4+ T cells, or CD137+/CD69+ events among CD8+ T cells, after subtracting the equivalent AIM+ frequency in the DMSO-stimulated control. **A)** Mann-Whitney U-test comparing AIM responses between patients receiving baseline prednisone (n = 17) and those not receiving prednisone (n = 16). Symbols differentiate between kidney (black circles), liver (red squares) and lung (blue triangles) transplant recipients. **B)** Comparisons of AIM responses between kidney (n = 14), liver (n = 11) and lung (n = 8) transplant recipients. Dunn’s multiple comparisons p-values are shown following a Kruskal-Wallis test. **C)** Semilogarithmic regression analysis of the relationship between age and T cell AIM responses, testing the null hypothesis of the slope being equal to zero.

Using semilogarithmic regression to model the effect of age on AIM responses, we identified significant declines in AIM responses with age (**Figure 4C**). The regression line slope was significantly different from zero for CD4+ ancestral-(y = -0.0108x + 0.0100, p = 0.0390) and BA.4/5-specific responses (y = -0.0127x + 0.0100, p = 0.0238), with even stronger relationships identified for CD8+ responses (ancestral, y = -0.0194x + 0.3949, p = 0.0146; BA.4/5, y = - 0.0267x + 0.6728, p = 0.0059). There were no significant differences in post-third dose AIM responses between males and females (**Figure S5A**), or between patients vaccinated primarily with Pfizer-BioNTech BNT162b2 compared to Moderna mRNA-1273 (**Figure S5B**).

### SARS-CoV-2-specific T cell responses correlate with circulating anti-Spike antibodies

We investigated correlations between CD4+ T cell AIM responses and previously published anti-RBD binding antibody titres measured in the same individuals at the same timepoints^9^. CD134+/CD69+ CD4+ T cell AIM responses correlated strongly with antibody levels (**Figure 5A**, **Table 2**). At pre- and post-second dose timepoints, there were weak to moderate correlations with antibody titres for CD4+ T cell responses to ancestral SARS-CoV-2 (pre-second dose Spearman’s r = 0.2369, p = 0.2884; post-second dose r = 0.3064, p = 0.0514) and BA.4/5 (pre-second dose r = 0.4066, p = 0.0604; post-second dose r = 0.3091, p = 0.0493). However, the strength and significance of these correlations increased over time, even without additional vaccinations, and were further enhanced by a third dose, for both responses to the ancestral strain (six months r = 0.8028, p = 0.0082; post-third dose r = 0.6755, p < 0.0001) and BA.4/5 (six months r = 0.6554, p = 0.0454; post-third dose r = 0.6411, p < 0.0001). By contrast, antibody correlations with CD8+ AIM responses did not follow a clear trend and were only significant for post-third dose ancestral strain responses (r = 0.3527, p = 0.0441) (**Figure 5A**, **Table 2**).

**Figure 5.**
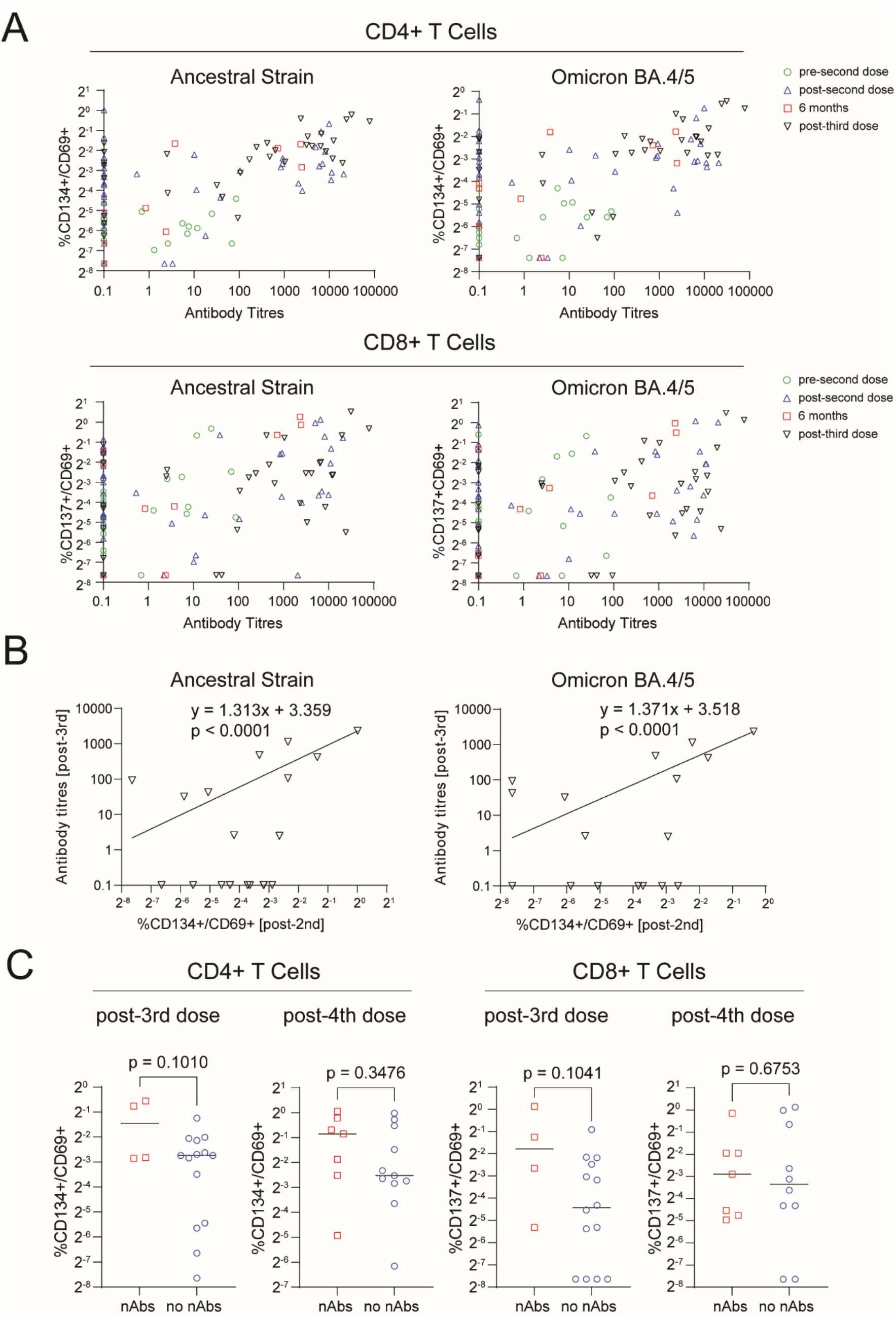
SARS-CoV-2-specific T cell activation-induced marker responses correlate with Spike-specific antibody titres in solid organ transplant recipients. Relationships of T cell activation-induced marker (AIM) responses with previously published anti-Spike binding and neutralizing antibodies are shown in solid organ transplant recipients^9^. PBMCs were collected from COVID-19-vaccinated SOTRs at pre-second dose, post-second dose, six months post-first dose and post-fourth dose timepoints. AIM responses are shown as frequencies of CD134+/CD69+ or CD137+/CD69+ events among CD4+ or CD8+ T cells, respectively, following stimulation for 20 h with 1 µg/mL SARS-CoV-2 ancestral strain or Omicron BA.4/5 Spike peptides, after subtracting the equivalent AIM+ frequency in the DMSO-stimulated control. **A)** Spearman’s correlations of CD4+ and CD8+ T cell activation-induced marker responses to ancestral or Omicron BA.4/5 variants of SARS-CoV-2 with circulating anti-Spike antibody titres. Correlations were analyzed separately at pre-second dose (green circles, n = 21), post-second dose (blue triangles, n = 40), six months post-first dose (red squares, n = 10) and third dose (black triangles, n = 33) vaccination timepoints. **B)** Log-log regression analysis of post-second dose CD4+ AIM responses and post-third dose anti-RBD antibody titres in SOTRs who had no detectable post-second dose antibody titres, testing the null hypothesis of slope being equal to zero. **C)** Comparison of post-third dose and post-fourth dose CD4+ and CD8+ T cell AIM responses to BA.4/5 in patients with (nAbs) or without (no nAbs) detectable BA.4/5 neutralization titres. Groups were compared by unpaired two-sample t-tests on log2-transformed AIM frequencies.

**Table 2.** Correlations of anti-RBD titres with T cell activation-induced marker responses.

| Timepoint | Variant | Spearman's r | P-value | Significance |
| --- | --- | --- | --- | --- |
| *CD4+ T cells (CD134+/CD69+)* |  |  |  |  |
| pre-2nd | Ancestral | 0.2369 | 0.2884 | ns |
| pre-2nd | BA.4/5 | 0.4066 | 0.0604 | ns |
| post-2nd | Ancestral | 0.3064 | 0.0514 | ns |
| post-2nd | BA.4/5 | 0.3091 | 0.0493 | * |
| 6 months | Ancestral | 0.8028 | 0.0082 | ** |
| 6 months | BA.4/5 | 0.6554 | 0.0454 | * |
| post-3rd | Ancestral | 0.6755 | <0.0001 | **** |
| post-3rd | BA.4/5 | 0.6411 | <0.0001 | **** |
| *CD8+ T cells (CD137+/CD69+)* |  |  |  |  |
| pre-2nd | Ancestral | 0.4273 | 0.0534 | ns |
| pre-2nd | BA.4/5 | 0.3288 | 0.1456 | ns |
| post-2nd | Ancestral | 0.2832 | 0.0727 | ns |
| post-2nd | BA.4/5 | 0.2726 | 0.0847 | ns |
| 6 months | Ancestral | 0.6457 | 0.0512 | ns |
| 6 months | BA.4/5 | 0.6241 | 0.0591 | ns |
| post-3rd | Ancestral | 0.3527 | 0.0441 | * |
| post-3rd | BA.4/5 | 0.3313 | 0.0596 | ns |

ns, non-significant; \*p <0.05, \*\*p <0.01; \*\*\*p <0.001; \*\*\*\*p <0.0001

As a substantial proportion of patients showed strong T cell responses but no detectable antibody titres post-second dose, we assessed whether post-second dose CD4+ T cell responses could predict subsequent post-third dose antibodies in these patients (n = 19). Log-log regression analysis identified a strong positive relationship, with patients who demonstrated stronger T cell responses to ancestral SARS-CoV-2 (y = 1.313x + 3.359, p < 0.0001) or BA.4/5 (y = 1.371x + 3.518, p < 0.0001) subsequently showing stronger post-third dose antibody responses (**Figure 5B**). We also assessed the relationship of post-third dose T cell AIM responses with the presence or absence of BA.4/5-specific nAbs in a subset of patients (n = 18). Although only four patients developed nAbs post-third dose, there was a clear trend toward higher CD4+ and CD8 AIM+ frequencies in these patients (**Figure 5C**). This trend was less apparent after a fourth dose.

### Significant escape from T cell recognition by the novel variant XBB.1.5 in SOTRs

We next asked whether three-dose ancestral COVID-19 vaccination induced T cells capable of cross-recognizing the novel XBB.1.5 variant, using whole protein isolates in a subset of the study cohort (n =10) for which we had additional pre-second dose and post-third dose PBMC samples. Although few XBB.1.5-specific T cells were observed prior to a second dose, three COVID-19 vaccine doses significantly induced XBB.1.5-responsive CD4+ T cells (**Figure 6A**). CD8+ AIM responses to XBB.1.5 were detected in only four of 10 patients after three doses. CD4+ CD134+/CD69+ AIM responses to XBB.1.5 were significantly weaker than responses to ancestral SARS-CoV-2 at pre-second dose (5.0-fold lower, p = 0.0078) and post-third dose timepoints (2.0-fold lower, p = 0.0020) (**Figure 6B**). CD8+ CD137+/CD69+ AIM responses to XBB.1.5 clearly trended lower (4.8-fold lower, p = 0.0781) at the pre-second dose timepoint, with XBB.1.5 responses detectable in only two of ten patients (seven of ten patients responded to the ancestral protein), and were significantly weaker than ancestral SARS-CoV-2 responses after three doses (18-fold lower, p = 0.0156) (**Figure 6B**).

**Figure 6.**
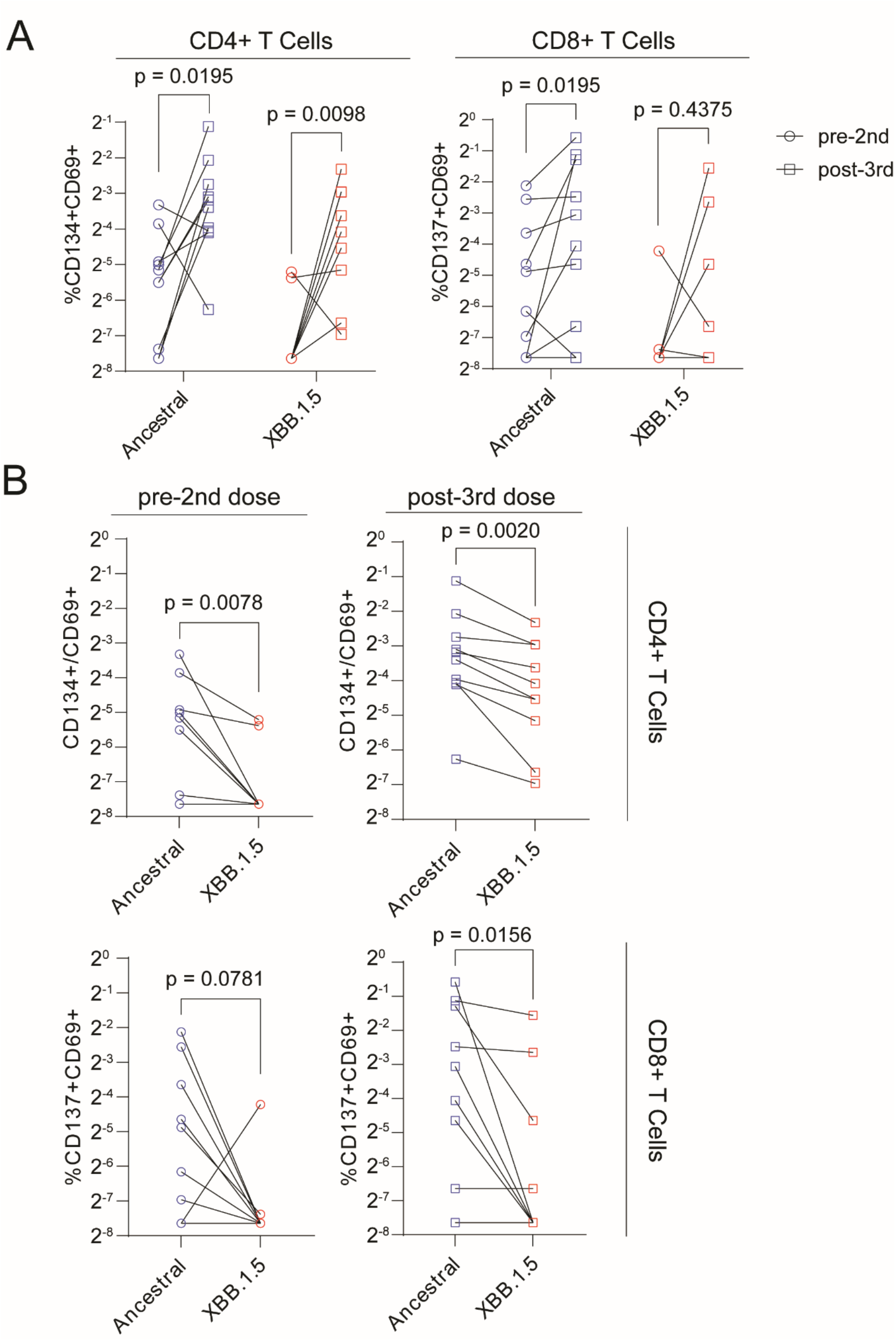
The SARS-CoV-2 variant XBB.1.5 partially escapes recognition by vaccine-induced T cells in solid organ transplant recipients. CD4+ and CD8+ T cell AIM responses to SARS-CoV-2 ancestral strain Spike protein and XBB1.5 Spike protein, measured prior to a second dose and after three doses of a COVID-19 mRNA vaccine in solid organ transplant recipients (n = 10). Following a 44-h stimulation of patient PBMCs with 0.5 µg/mL SARS-CoV-2 ancestral strain or XBB.1.5 Spike protein, CD4+ T cell activation-induced marker (AIM) responses were quantified as frequencies of CD134+/CD69+ events among CD4+ T cells, while CD8+ AIM responses were measured as frequencies of CD137+/CD69+ events among CD8+ T cells, after subtraction of the equivalent AIM+ frequency in the PBS-stimulated control. Paired Wilcoxon signed-rank test p-values are shown. **A)** Comparisons of AIM responses between pre-second dose and post-third dose timepoints. **B)** Comparisons of AIM responses to ancestral SARS-CoV-2 and XBB.1.5.

## Discussion

As SARS-CoV-2 variants continue to evolve, understanding vaccine immunogenicity in vulnerable populations is critical to informing vaccination strategies. SOTRs show impaired humoral and cellular responses to mRNA vaccines^5,9,28,30^, corresponding with increased susceptibility to infection^4,25^. In the present study, we have provided the first AIM based-characterization of COVID-19 vaccine-induced T cell responses in SOTRs. We demonstrate significant T cell responses to vaccination which persist over time and are enhanced by booster doses. Although monovalent vaccination with the ancestral strain induced T cells capable of cross-recognizing BA.4/5 and XBB.1.5, responses to these variants were weaker than those to the ancestral strain.

We observed strong induction of SARS-CoV-2-specific AIM responses to both ancestral SARS-CoV-2 and BA.4/5 by second and third doses of mRNA vaccine, particularly among CD4+ T cells. Although responses trended lower over time at the six-month and one-year timepoints, CD4+ T cell responses remained significantly elevated above pre-second dose levels and were further enhanced by a fourth dose. Our data confirm studies showing that SOTRs mount significant T cell responses to mRNA vaccination^9,25,28,29,31^, and represent the first AIM-based characterization of BA.4/5-cross-reactive T cells in SOTRs. We show that even two-dose mRNA vaccination significantly induces BA.4/5 cross-reactivity, however, this declines over time and at least one additional (i.e. third) dose may be required to achieve lasting BA.4/5-specific T cell memory. Indeed, longitudinal analyses of immune-suppressed patients with diverse pathologies observe sharp declines in post-second dose responses^28,50^, while responses remain stable at similar magnitudes to healthy individuals following third and fourth doses^50^. Interestingly, breakthrough infection did not appear to further enhance BA.4/5-specific responses in patients who received a fourth dose, despite cases occurring during a global Omicron wave. Of note, these infections occurred more than four months (IQR 111-148 days) prior to one-year sampling, thus the acute T cell response to infection was likely not captured.

We also established clinical factors associated with BA.4/5-specific T cell responses to vaccination. Although responses in patients taking prednisone were decreased, consistent with studies of BA.4/5 neutralization in SOTRs^9^, all lung recipients in our cohort, and no liver recipients, were receiving prednisone in accordance with regional guidelines^51^.When analyzing differences between organ groups, lung recipients responded weakly compared to liver and kidney recipients. This is likely due to the more intense immunosuppression given to lung transplant recipients^52,53^. Others have demonstrated impaired vaccine-induced IFN-γ responses and increased COVID-19 mortality in lung recipients^5,54,55^. We also demonstrate a negative relationship between age and BA.4/5-specific AIM responses. CD8+ AIM responses showed particularly strong declines with increasing patient age, congruent with greater susceptibility to ageing in CD8+ T cells than CD4+ T cells^56–58^.

As CD4+ T cells promote high-affinity humoral responses and B cell memory^59^, we analyzed correlations of BA.4/5-specific AIM responses with anti-RBD and nAb titres^9^. Correlations between CD4+ AIM responses and anti-RBD titres were non-significant prior to a second dose, but increased in strength and significance with subsequent vaccinations, and over time even without additional vaccinations. At the six-month timepoint or after three doses, patients who developed antibody responses generally showed strong CD4+ AIM responses, suggesting that a third dose is necessary to optimally stimulate both immune compartments and that the relationship between cellular and humoral immunity may also strengthen with time. However, a substantial number of patients with no detectable anti-RBD titres after two or three doses showed strong AIM responses, hinting that T cells could play a critical protective role in these antibody-deficient SOTRs^21,60^. Furthermore, post-second dose T cell responses strongly predicted post-third dose antibody responses, supporting a causative relationship between antigen-specific CD4+ T cells and humoral responses. We also observed trending associations between BA.4/5-specific AIM responses and BA.4/5-nAbs, but these were non-significant as few patients developed post-third dose BA.4/5-nAbs.

The BA.4/5 and XBB.1.5 variants escape antibody-mediated neutralization in individuals vaccinated against the ancestral strain^13,16–20,61^. CD4+ and CD8+ T cell responses have thus far shown conserved cross-reactivity with Omicron^18,26,27,62^ including BA.4/5 in immune-compromised patients and SOTRs^31,63^, but no data existed for T cell responses to XBB.1.5. In contrast to previous research, we found significant decreases in T cell recognition of BA.4/5 relative to the ancestral strain. We are the first to investigate BA.4/5 cross-recognition in SOTRs using an AIM assay, which detects cytokine-negative T cells and rare populations with higher sensitivity than ICS^33,34^, lending itself well to detecting small differences. We obtained similar results for XBB.1.5, showing strongly impaired T cell cross-recognition in monovalent ancestral-vaccinated SOTRs. However, despite their lower magnitude, CD4+ responses to BA.4/5 and XBB.1.5 were induced by vaccination, consistent with broad epitope specificity of CD4+ T cells^64^, and displayed similar kinetics to ancestral responses. Thus, impairments in T cell cross-recognition may not severely impact vaccine immunogenicity for these variants. Large-scale epidemiological data in SOTRs show significantly decreased incidence of hospitalization and death following the emergence of Omicron, suggesting adequate vaccine-mediated protection from severe outcomes in this highly transmissible but low-virulence variant^4^. The full impact of the recent wave of XBB.1.5 infections on SOTRs remains to be seen.

Our study entails several inherent limitations. First, we are unable to draw conclusions regarding the magnitude of AIM responses in SOTRs relative to the general population as we did not include healthy controls in the study. This also precludes the extension of novel findings, such as impaired T cell responses to BA.4/5 and XBB.1.5, beyond the SOTR population. Second, our sample size was insufficient to adequately assess effects of certain clinical parameters such as antimetabolite or tacrolimus use, or whether AIM responses predict subsequent infection or hospitalization. Finally, our method provided more robust characterization of CD4+ T cell responses than CD8+ T cell responses. This may be a feature of the peptide mixes used, which consist of 15-mers. CD4+ T cells recognize peptides from 11-20 amino acids, while CD8+ T cells optimally recognize peptides of 8-11 amino acids^65^. However, CD8+ T cell responses were also poorly detected using whole protein stimulation (**Figure 6**), and others have observed lower magnitudes of Spike-specific CD8+ responses using diverse methods^43,44,60,66,67^. Thus, CD8+ T cells may preferentially respond to non-Spike antigens, including internal viral proteins^44^, and Spike-based assays may not capture the full complement of SARS-CoV-2-specific CD8+ T cells.

Overall, we demonstrate that SOTRs mount significant BA.4/5-cross-reactive T cell responses to ancestral COVID-19 mRNA vaccines, with booster doses enhancing responses. However, using a sensitive AIM assay, we show that cross-recognition of BA.4/5 and XBB.1.5 is significantly impaired in SOTRs. We also demonstrate strong correlations between CD4+ T cell responses and antibody responses, and identify weaker cellular responses in older adults and lung recipients receiving prednisone. Our data provide unique insights into the kinetics of variant-specific T cell responses in immune-compromised patients, with important implications for clinical and public health guidelines.

## Supporting information

Supplemental Digital Material

## Data Availability

All data produced in the present study are available upon reasonable request to the authors

## AUTHORSHIP PAGE

### Author Contributions

**TH** research design, paper writing, performance of the research, data analysis, statistical analyses

**SI** research design, paper writing, data analysis

**QH** research design, performance of the research

**GL** performance of the research, data analysis

**DY** performance of the research, data analysis

**DM** research design, performance of the research, paper writing

**SS** research design, paper writing

**VHF** data analysis, research design

**DK** research design

**SB** research design, performance of the research

**MKL** research design, paper writing

### Disclosures

D.K. has received consulting fees from Roche, GSK, Exevir and clinical trials grants from GSK. S. B. has received consultant fees from AstraZeneca and GSK, honoraria from Merck and research funding from Takeda. The remaining authors have no relevant conflicts to disclose.

### Funding

This work was supported by a grant from Public Health Agency of Canada / COVID-19 Immunity Task Force (AWD-019932 PHACA 2021) and the Canadian Institutes of Health Research (HUI-159423). MKL holds a salary award from the BC Children’s Hospital Research Institute and a Tier 1 Canada Research Chair in Engineered Immune Tolerance. This study was coordinated by the Canadian Donation and Transplantation Research Program (CDTRP).

## Abbreviations

AIM: activation-induced marker
BA.4/5: Omicron BA.4 and BA.5 variants
COVID-19: coronavirus disease of 2019
DMSO: dimethyl sulfoxide
ICS: intracellular cytokine staining
IQR: interquartile range
IFN-γ: interferon-γ
IL-2: interleukin-2
mRNA: messenger RNA
nAb: neutralizing antibody
PBMCs: peripheral blood mononuclear cells
RBD: receptor-binding domain
SARS-CoV-2: severe acute respiratory syndrome coronavirus 2
SOTR: solid organ transplant recipient

## Acknowledgements

We wish to express our deepest gratitude to those who participated in this study. Finally, we wish to thank Lisa Xu at the BC Children’s Hospital Research Institute Flow Cytometry Core for her invaluable assistance with data acquisition. The study was coordinated by the Canadian Donation and Transplantation Research Program.

## PREVenT COVID group investigators

PREVenT COVID group investigators: Jean-Sébastien Delisle (Hôpital Maisonneuve Rosemont (HMR), Sasan Hosseini-Moghaddam (University Health Network (UHN)), Dr. Héloïse Cardinal (Centre hospitalier de l’Université de Montréal (CHUM)), Dr. Mélanie Dieudé (Centre hospitalier de l’Université de Montréal (CHUM)), Dr. Normand Racine (Institut de Cardiologie de Montréal (ICM)), Dr. Karina Top (Dalhousie University), Dr. Gaston DeSerres (INSPQ, Public Health), Dr. Lori West (University of Alberta (UofA), CDTRP)), Dr. Marc Cloutier (HémaQuébec), Dr. Renée Bazin (Héma-Québec), Dr. Christopher Lemieux (Université Laval), Dr. Sacha De Serres (Université Laval), Dr. Atul Humar (University Health Network (UHN)), Dr. Sarah Shalhoub (London Health Sciences Center (LHSC)), Dr. Dima Kabbani (University of Alberta (UofA)), Dr. Marie-Josée Hébert (Centre hospitalier de l’Université de Montréal (CHUM)), Dr. Patricia Gongal (CDTRP), Kristian Stephens (CDTRP).

## PREVenT COVID Group Coordinators

Dr. Julie Turgeon (CHUM), Zineb Khrifi (CHUM), France Samson (Université Laval), Maryse Desjardins (ICM), Hélène Brown (ICM), Johanne Doiron (ICM), Cadence Baker (London Health Sciences Centre, LHSC), Taylor Toth (LHSC), Grant Luke (LHSC), Natalia Pinzon (UHN), Victoria G. Hall (UHN), Kimberly Robertson (UofA), Heather Mangan (UofA).

## Notes

### Author Declarations

This study of cell-mediated immunity was conducted at the University of British Columbia (UBC) with approval from the UBC Research Ethics Board (H21-01269).

