## Supplemental Digital Material for "SARS-CoV-2 variants Omicron BA.4/5 and XBB.1.5 significantly escape T cell recognition in solid organ transplant recipients vaccinated against the ancestral strain"

### Extended Methods

*Study design and enrollment*

The PREVenT-COVID study is a Canadian prospective study of vaccine immunogenicity in SOTRs across seven tertiary care transplant centres[9]. This study of cell-mediated immunity was conducted at the University of British Columbia (UBC) with approval from the UBC Research Ethics Board (H21-01269). All participants provided informed consent. Adult outpatient SOTRs at least 1 month post-transplant and taking at least one immunosuppressive therapeutic were enrolled at first dose of an approved COVID-19 mRNA vaccine, Pfizer-BioNTech BNT162b2 or Moderna mRNA-1273, beginning January 2021. Patients who reported a febrile illness in the past week, active cytomegalovirus viremia, use of rituximab in the past 6 months, or current intravenous immunoglobulin or acute rejection therapy were excluded. Whole blood was drawn from participants within 2 weeks prior to the 2$^{nd}$ vaccine dose, 3-6 weeks post-1$^{st}$ dose, 6 months post-1$^{st}$ dose, 3-6 weeks post-3$^{rd}$ dose and 1 year post-1$^{st}$ dose. 1-year samples served as post-fourth dose (3-6 weeks) samples in a subset of patients. PBMCs were collected from blood samples and cryopreserved at -80ºC. Participants were to inform the study team if they developed COVID-19, for confirmation via PCR or rapid antigen test.

*SARS-CoV-2 ancestral and Omicron BA.4/5 Spike peptides*
SARS-CoV-2 ancestral strain (PM-WCPV-S-1, JPT) and Omicron BA.4/5 (PM-SARS2-SMUT10-1, JPT) peptide pools (15-mers overlapping by four amino acids) corresponding to the complete Spike proteins were aliquoted in 30% DMSO and stored at -80ºC.

*SARS-CoV-2 ancestral and XBB.1.5 Spike proteins*
The HexaPro expression plasmid was a gift from Jason McLellan (Addgene plasmid #154754; http://n2t.net/addgene:154754; RRID: Addgene_154754) which was used to construct the D614G mutant via site-directed mutagenesis (Q5 Site-Directed Mutagenesis Kit, New England Biolabs). The XBB.1.5 HexaPro S protein gene was synthesized and inserted into pcDNA3.1 (GeneArt Gene Synthesis, Thermo Fisher Scientific). Expi293F cells (Thermo Fisher, Cat# A14527) were grown in suspension culture using Expi293 Expression Medium (Thermo Fisher, Cat# A1435102) at 37 °C, 8% CO2. Cells were transiently transfected at a density of 3 × 106 cells/mL using linear polyethylenimine (Polysciences Cat# 23966-1). The media was supplemented 24 h after transfection with 2.2 mM valproic acid, and expression was carried out for 3–7 days at 37 °C, 8% CO2.

To purify spike protein ectodomains, supernatant was loaded onto a 5 mL HisTrap excel column (Cytiva). The column was washed for 20 column volumes (CVs) with wash buffer (20 mM Tris pH 8.0, 500 mM NaCl), 5 CVs of wash buffer supplemented with 20 mM imidazole, and the protein eluted with elution buffer (20 mM Tris pH 8.0, 500 mM NaCl, 500 mM imidazole). Elution fractions containing the protein were pooled and concentrated (Amicon Ultra 100 kDa cut off, Millipore Sigma) for gel filtration. Gel filtration was conducted using a Superose 6 10/300 GL column (Cytiva) pre-equilibrated with GF buffer (20 mM Tris pH 8.0, 150 mM NaCl). Peak fractions corresponding to soluble protein were pooled and concentrated (Amicon Ultra 100 kDa cut off, Millipore Sigma) Protein samples were flash-frozen in liquid nitrogen and stored at −80°C.

*Activation-induced marker assays*

Cryopreserved PBMCs were thawed into 37ºC Immunocult XF (10981, STEMCELL) with 1% penicillin/streptomycin and 50 U/mL benzonase (70664-3, Novagen). PBMCs were rested overnight at 37ºC in Immunocult XF with 1% penicillin/streptomycin. For ancestral and BA.4/5 peptide-based assays, PBMCs were stimulated for 20 h at 4-5x$10^5$ cells per condition in 200 uL wells of a 96-well U-bottom plate with 1 µg/mL SARS-CoV-2 ancestral strain or Omicron BA.4/5 peptides. Cytostim (0.05%) (130-092-172, Miltenyi) and equimolar (0.12%) DMSO were used as positive and negative controls, respectively, and an influenza vaccine (2.5%) (Fluzone® Quadrivalent, Sanofi-Pasteur) stimulation was used as an assay control. For ancestral and XBB.1.5 protein-based assays, stimulation was performed with whole protein isolates at 0.5 µg/mL for 44 h, using an equivolumetric PBS as a negative control, 1 µg/mL ancestral strain peptides as an assay control and 0.05% Cytostim as a positive control.

CD4 and CD107a staining antibodies were added during stimulation, while surface staining for CD3, CD4, CD8, CD25, CD69, CD134, and CD137 was performed after the stimulation period. Data were acquired on an LSR Fortessa (BD) flow cytometer on high-throughput sampler (HTS) mode. Antigen-specific CD4+ T cells were defined as CD134+/CD25+, CD134+/CD69+ or CD137+/CD69+ while antigen-specific CD8+ T cells were defined as CD107a+/CD69+, CD107a+/CD137+ or CD137+/CD69+, using widely reported markers[23,36–44].

*Data Analysis*

Data were analyzed using FlowJo v10.8.1 and GraphPad Prism v10.0.0. Antigen-specific T cells were quantified as the percentage of CD4+ or CD8+ T cells expressing each AIM marker, after subtracting the equivalent frequency of AIM+ cells in the unstimulated condition. AIM frequencies less than 0.005%, considered the limit of detection for the assay, were set to equal 0.005% to avoid negative or zero values. Samples with fewer than 10 000 total CD4+ or CD8+ cells were excluded from analyses. AIM responses were compared across timepoints using a mixed-effects analysis (REML) on $\log_2$-transformed data with the assumption of sphericity, followed by post-hoc Dunnett's multiple comparisons test. Patients who received a fourth dose or contracted COVID-19 were excluded from this analysis.

Wilcoxon signed-rank tests were used to compare 1-year to post-3rd dose responses between post-fourth dose (within 3-6 weeks), control (neither received fourth dose nor contracted COVID-19) and hybrid (fourth dose and contracted COVID-19) groups. Changes from post-third dose between groups were compared using a Kruskal-Wallis test with Dunn's multiple comparisons. Responses to the ancestral strain and Omicron BA.4/5, as well as responses to the ancestral strain and XBB.1.5, were compared by Wilcoxon signed-rank test on post-3rd dose AIM+ frequencies. Median post-3rd dose AIM responses between groups defined by clinical parameters were compared using Mann-Whitney and Kruskal-Wallis (with Dunn's multiple comparisons) tests as appropriate. Semilogarithmic regression analysis was performed to model the continuous effects of age and time since transplantation on post-3rd dose AIM responses, testing the null hypothesis of slope being equal to zero.

Spearman's correlations were used to assess relationships between post-third dose T cell AIM responses and previously published serum anti-RBD titres from the same individuals[9]. In patients with undetectable anti-RBD antibodies post-second dose, log-log regression was used to assess the relationship between post-second dose T cell AIM responses and subsequent development of post-third dose antibody titres. Post-third dose AIM responses to BA.4/5 were also compared between individuals with and without detectable BA.4/5 neutralization titres[9].

#### Supplemental Figures

### Figure S1

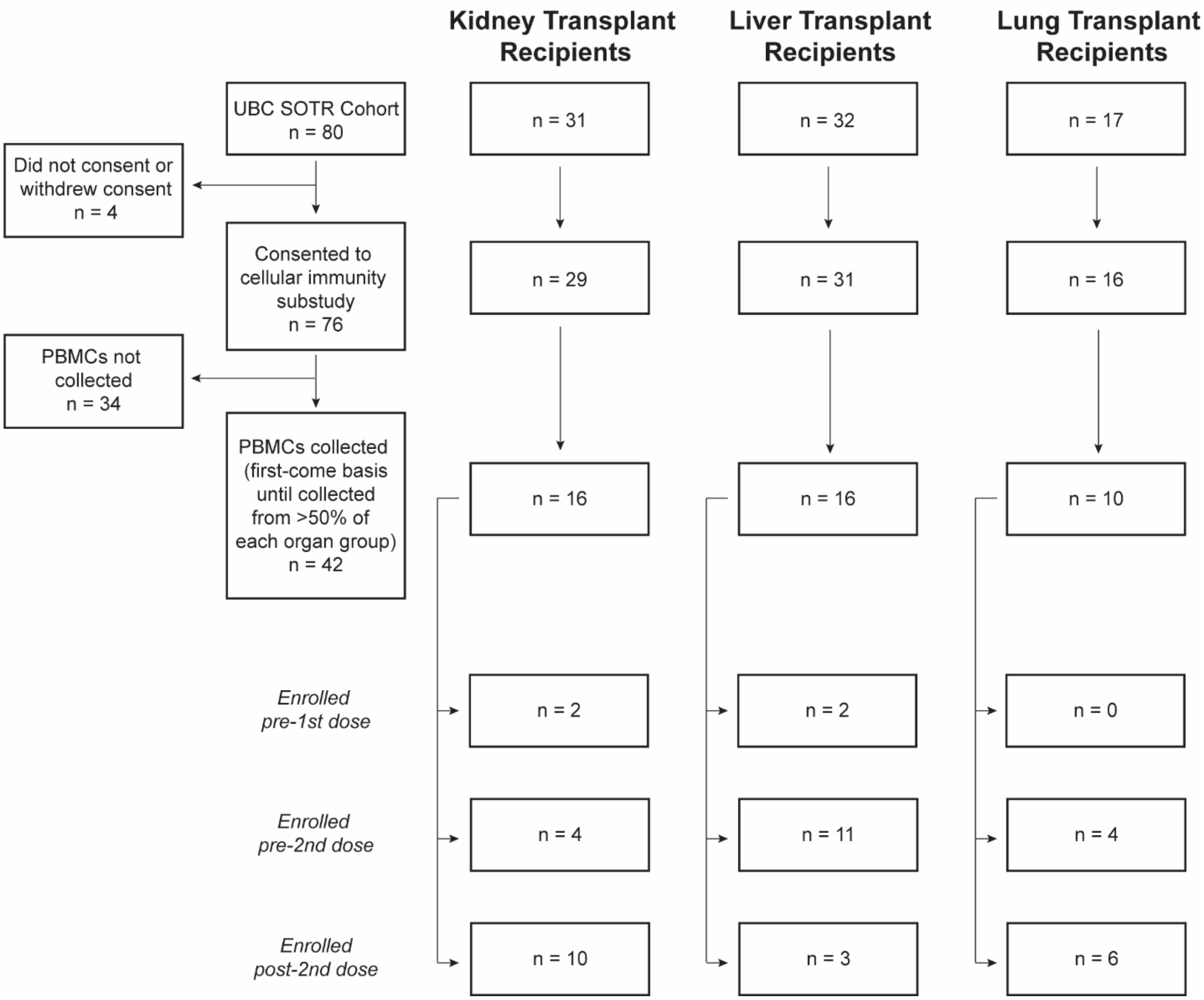


**Figure S1. Cohort overview.** Solid-organ transplant recipients (SOTRs) were recruited for the cellular immunity substudy from the University of British Columbia (UBC) PREVenT cohort (n = 80). Four SOTRs did not consent or withdrew consent. The remaining SOTRs were enrolled (PBMCs collected) at pre-second dose, post-second dose or post-third dose timepoints until PBMCs from over half of each organ group (kidney, liver or lung recipients) had been collected. The final sample size (n = 42) thus included 16 kidney recipients, 16 liver recipients and 10 lung recipients.

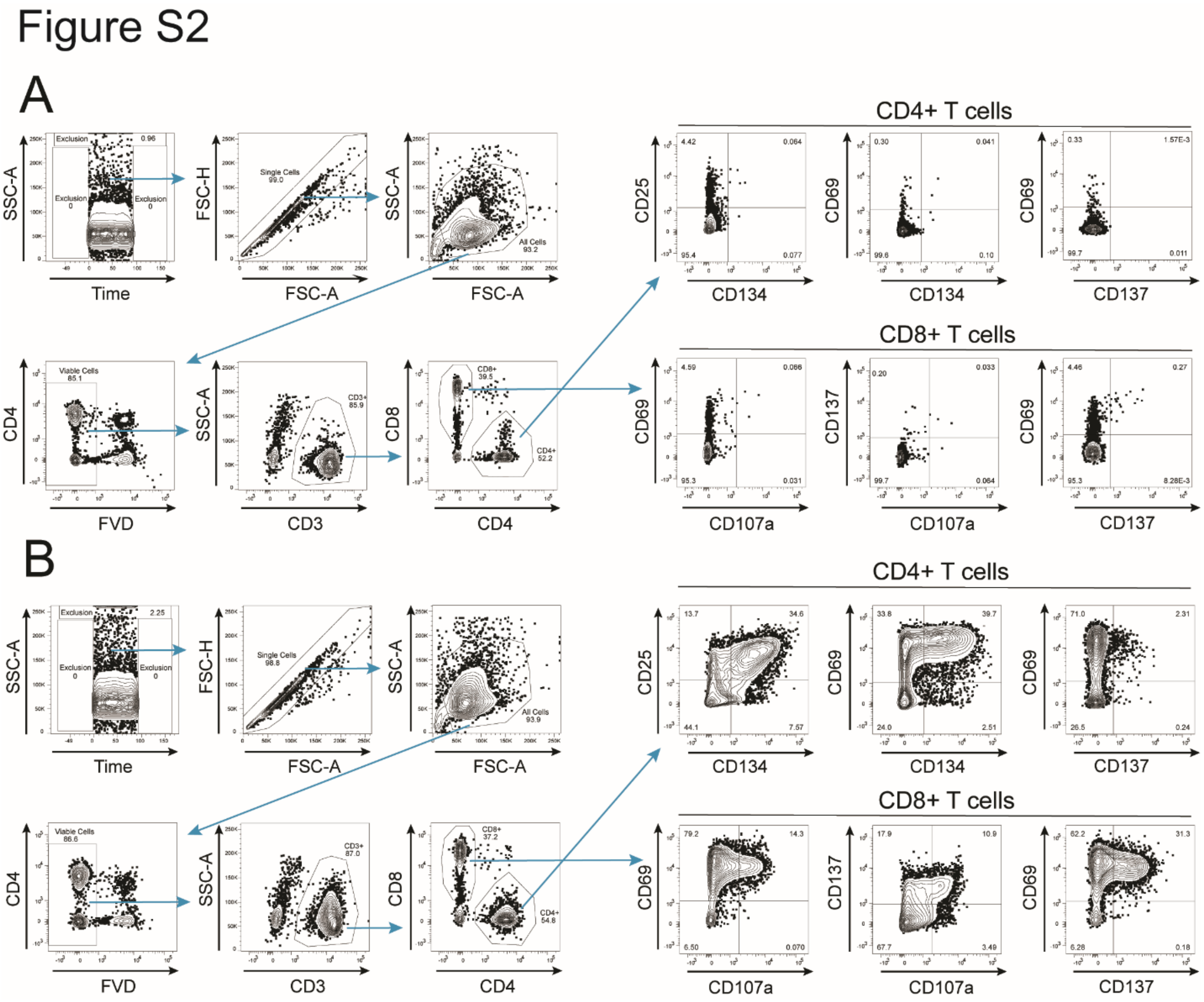


**Figure S2. Representative gating strategy.** Example flow cytometry data are shown for PBMCs from SOTRs following a 20 h stimulation with **A)** DMSO (0.12%) and **B)** Cytostim (0.05%). Activation-induced marker (AIM)+ cell frequencies were measured within live CD4+ or CD8+ single cell populations. AIM+ CD4+ T cells were defined as co-expressing CD134 and CD25, CD134 and CD69 or CD137 and CD69. AIM+ CD8+ T cells were defined as co-expressing CD107a and CD69, CD107a and CD137 or CD137 and CD69. The Cytostim-stimulated positive control for each sample was used to define the respective AIM+ population for each AIM.

### Figure S3

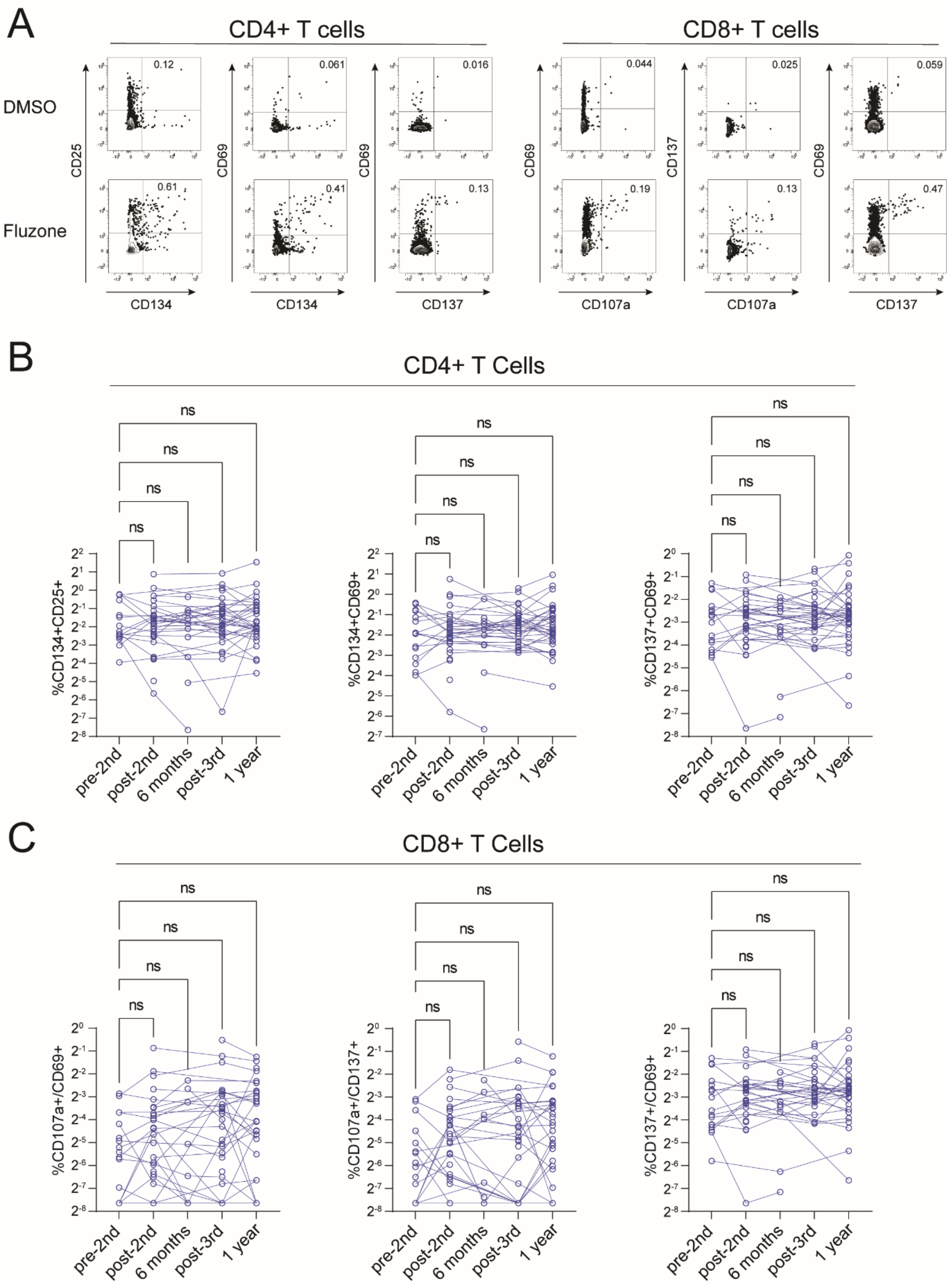

A
CD4+ T cells
CD8+ T cells
DMSO
Fluzone
CD25
CD69
CD69
CD69
CD137
CD69
CD134
CD134
CD137
CD107a
CD107a
CD137
0.12
0.061
0.016
0.044
0.025
0.059
0.61
0.41
0.13
0.19
0.13
0.47
B
CD4+ T Cells
%CD134+CD25+
%CD134+CD69+
%CD137+CD69+
ns
pre-2nd
post-2nd
6 months
post-3rd
1 year
C
CD8+ T Cells
%CD107a+/CD69+
%CD107a+/CD137+
%CD137+/CD69+

**Figure S3. The activation-induced marker assay is specific to SARS-CoV-2.** PBMCs from solid organ transplant recipients (SOTRs, n = 42) vaccinated with COVID-19 mRNA vaccines were stimulated with 2.5% Fluzone® Quadrivalent influenza vaccine (Sanofi-Pasteur) as an irrelevant antigen control. **A)** Representative flow cytometry data showing CD4+ and CD8+ T cell activation-induced marker (AIM) responses. Influenza-specific CD4+ **(B)** and CD8+ **(C)** T cell activation-induced marker responses at pre-second dose, post-second dose, six months post-first dose, post-third dose and one year post-first dose throughout the COVID-19 vaccination schedule. All data are shown as AIM+ frequencies after subtracting the equivalent AIM+ frequency in the DMSO-stimulated control. SOTRs who received a fourth dose or contracted COVID-19 before the one year timepoint were included.

### Figure S4

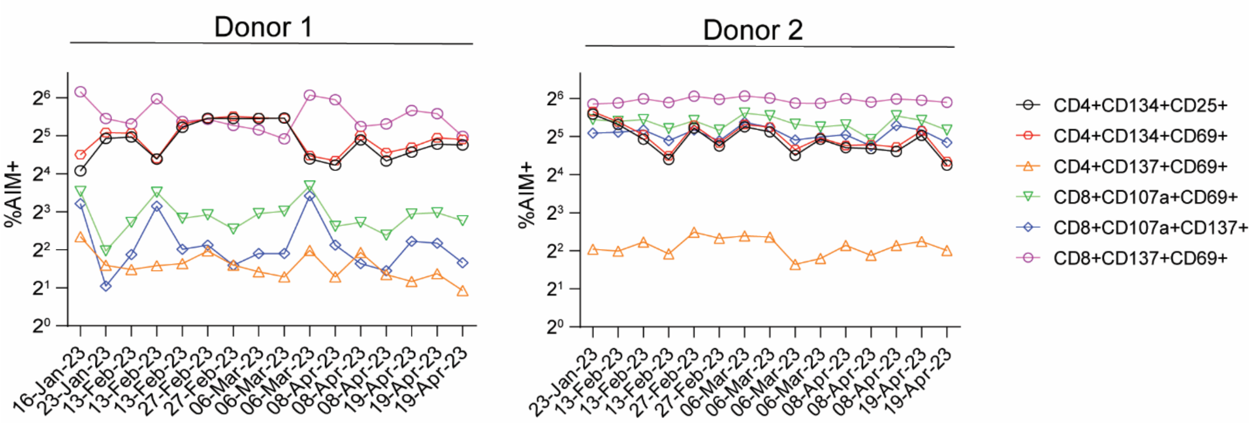


**Figure S4. The activation-induced marker assay demonstrates low technical variability over time.** Representative PBMCs from two healthy donors used as batch controls display consistent activation-induced marker responses over time when stimulated with 0.05% Cytostim (anti-CD3/CD28) (Miltenyi).

### Figure S5

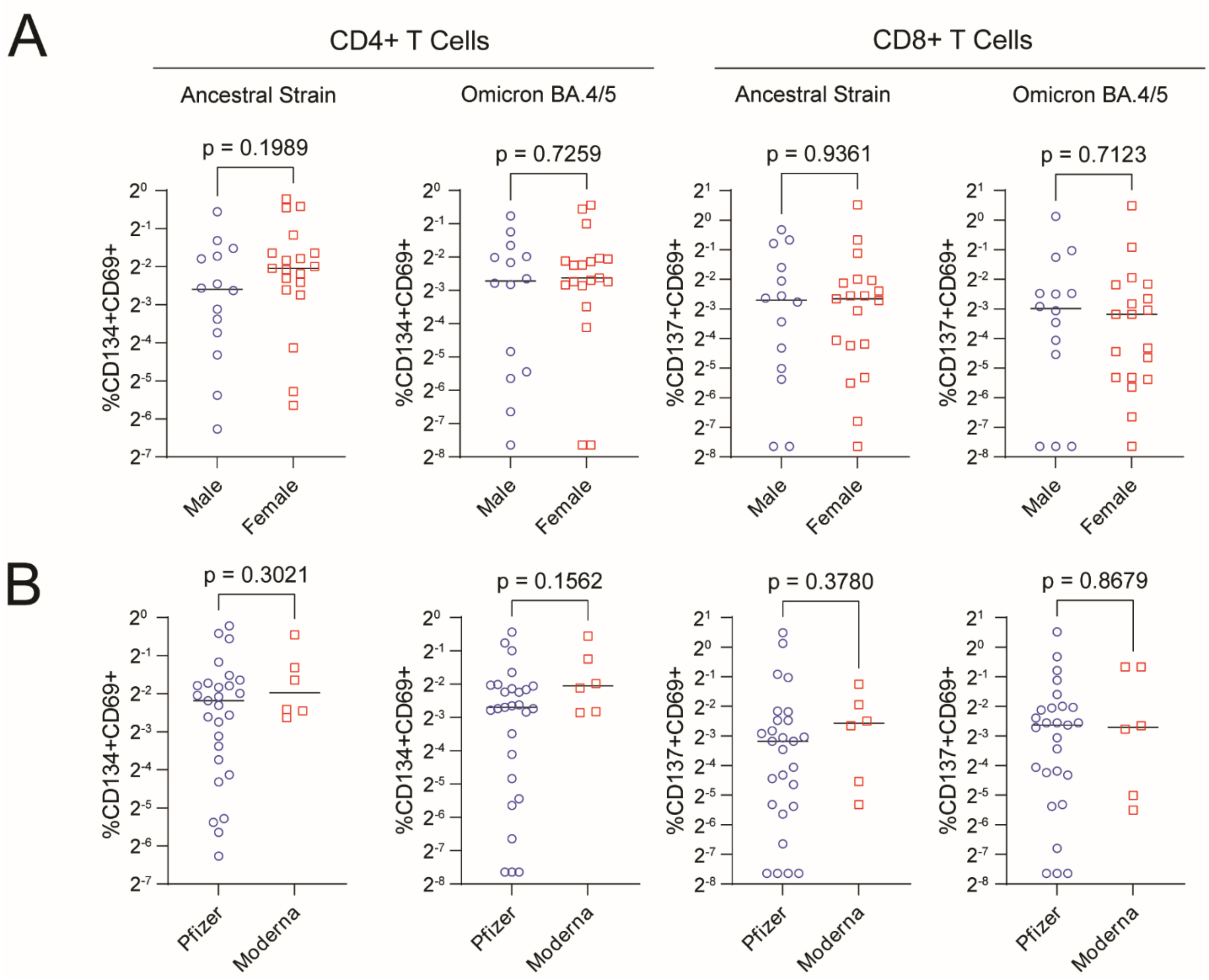


**Figure S5. Relationships of T cell activation-induced marker responses with sex and vaccine type in solid organ transplant recipients.** Post-third dose PBMCs from solid organ transplant recipients vaccinated with COVID-19 mRNA vaccines were stimulated with 1 µg/mL SARS-CoV-2 ancestral or Omicron BA.4/5 Spike peptides for 20 h. Activation-induced marker (AIM) responses were quantified by flow cytometry as frequencies of CD134+/CD69+ or CD137+/CD69+ events among CD4+ and CD8+ T cells, respectively, after subtracting the equivalent frequency in the DMSO-stimulated control. **A)** AIM responses in male (blue circles, n = 14) and female (red squares, n = 19) patients were compared using a Mann-Whitney U-test. **B)** AIM responses in patients who had received Pfizer-BioNTech BNT162b2 (Pfizer, blue circles, n = 27) or Moderna mRNA-1273 (Moderna, red squares, n = 6) vaccines for at least two of three doses were compared using a Mann-Whitney U-test.
